# Genus-level transfer learning of Matrix-assisted laser desorption/ionization time-of-flight mass spectrometry data predicts antibiotic resistance with greater accuracy

**DOI:** 10.1101/2025.04.25.25326423

**Authors:** Ming-I Chen, Yin-Tzer Shih, Ying-Lin Hsu, Yi-Hsin Chen

**Affiliations:** Doctoral Program in Big Data Analytics for Industrial Applications, National Chung Hsing University, Taichung, Taiwan; Department of Applied Mathematics and Institute of Statistics, National Chung Hsing University, Taichung, Taiwan; Department of Nephrology, Taichung Tzu Chi Hospital, Taichung, Taiwan; School of Medicine, Tzu Chi University, Hualien, Taiwan

**Keywords:** MALDI-TOF MS, antimicrobial resistance, transfer learning

## Abstract

Bacterial resistance, driven by excessive antibiotic use, has rendered many traditional antibiotics ineffective. Despite the advantages of applying matrix-assisted laser desorption/ionization time-of-flight mass spectrometry (MALDI-TOF MS) to predict bacterial antimicrobial resistance, limited databases and a lack of high-quality data hinder this effort. This study aimed to address this lack of data by integrating MALDI-TOF MS technology with transfer learning approaches, utilizing genus-level data, to predict species-specific bacterial antimicrobial resistance. The data were retrieved from the DRIAMS dataset. A multilayer perceptron deep neural network was applied for pretraining and fine-tuning, integrating genus-level data to enhance the accuracy of the species-specific predictions. Notably, fine-tuned models enhanced antibiotic resistance prediction. In *Staphylococcus aureus*, the area under the receiver operating characteristic curve (AUROC) improved from 0.95 to 0.96 and the area under the precision-recall curve (AUPRC) improved from 0.85 to 0.88 for oxacillin; for fusidic acid, the AUROC improved from 0.80 to 0.81 and the AUPRC from 0.32 to 0.36; and for ciprofloxacin, the AUROC improved from 0.80 to 0.82 while the AUPRC remained 0.57. In *Klebsiella pneumoniae*, the AUROC held steady at 0.75, while the AUPRC declined from 0.45 to 0.41 for ciprofloxacin, indicating potential limitations for certain pathogen-antibiotic combinations. Despite such variability, the overall improvements across multiple strains highlight the potential of transfer learning in antimicrobial resistance prediction and underscore the need to expand genus-level databases to improve species-level diagnostic accuracy, thereby enhancing clinical diagnostics.

## Introduction

Owing to multidrug resistance from the widespread misuse and overuse of antibiotics, the efficacy of conventional antimicrobial therapies against many bacterial species has been significantly compromised, thereby exacerbating challenges of infection management [1]. This situation increases treatment costs, exacerbates patient suffering, and increases mortality rates [2]. The combination of prolonged and inappropriate antibiotic prescribing practices [3, 4], combined with antibiotic contamination in water and soil as well as antibiotic use in agriculture, have accelerated the spread of resistance genes [5]. Consequently, antimicrobial resistance (AMR) has emerged as an increasingly urgent global public health crisis [6]. According to the United Nations Environment Programme, in 2019 alone, drug-resistant infections and bacterial AMR were directly responsible for approximately 1.27 million and 4.95 million mortalities worldwide, respectively. Alarmingly, by 2050, annual AMR-related deaths could rise to 10 million comparable to the global cancer mortality rate reported in 2020.

The continuous development of novel diagnostic methods for the rapid identification of pathogenic microorganisms is imperative for providing more effective treatment options. The matrix-assisted laser desorption/ionization time-of-flight mass spectrometry (MALDI-TOF MS) system effectively identifies pathogens and may provide species-specific mass spectral patterns of microbial proteins for the rapid identification of species [7]. MALDI-TOF MS demonstrates notably higher accuracy and speed than traditional gel-based protein or DNA fingerprinting techniques, and has thus rapidly become a standard technology in clinical microbiology laboratories [8]. Compared with traditional culturing, MALDI-TOF MS reduces the microbiological diagnosis period to within 24 h [9]. However, its capability to identify viruses is limited; their unique biological characteristics require the optimization of operational parameters [10]. MALDI-TOF MS presents broad potential in identifying various microorganisms. However, specific conditions, such as the homogeneity of infection and sufficient cell count—are critical for ensuring result reliability [11]. Despite advancements, additional challenges—including method reproducibility and data storage—continue to hinder the clinical application of MALDI-MS-based imaging proteomics [12]. Reportedly, qShot MALDI analysis, a new method, offers a cost-effective approach for quantitatively analyzing phospholipids, further expanding the potential applications of MALDI-TOF MS in clinical diagnostics [13]. However, the lack of comprehensive databases and high-quality data continues to limit its widespread application. Hence, establishing and expanding more robust and standardized databases is necessary for enhancing the application scope and precision of MALDI-TOF MS [14,15].

Transfer learning—a machine learning technique—repurposes a previously trained model for a secondary task. This approach capitalizes on information from large datasets to enhance the performance of models working with limited data [16]. Transfer learning has been applied across various medical fields, including medical image analysis and disease prediction, to address the limitations of MALDI-TOF MS and the data scarcity associated with species-specific datasets [17,18]. For example, Seddiki et al. explored transfer learning and cumulative representation learning using a one-dimensional convolutional neural network (1D-CNN) for MS data classification. They addressed challenges related to small sample sizes by leveraging various small datasets across different biological contexts, which significantly improved classification accuracy [19]. Moreover, by leveraging models pretrained on extensive datasets, transfer learning can improve diagnostic and predictive accuracy [20,21]. Hamid and Friedberg [22] investigated the application of transfer learning in the context of protein sequence data. Their method utilized neural networks pretrained on the UniRef50 database and demonstrated excellent adaptability and predictive capabilities, even with small-scale, low-redundancy datasets. Supporting this, Wang et al. [23] developed a CNN model that accurately predicts vancomycin-resistant *Enterococcus faecium* by analyzing complete mass spectrum profiles. Furthermore, using E. coli genomic and antimicrobial susceptibility data produced in their own laboratory, Ren et al. [24] reinforced these findings by demonstrating that deep transfer learning can achieve highly accurate predictions of AMR for novel antibiotics. Additionally, López-Cortés et al. employed a deep learning approach (1D-CNN) to directly analyze raw Mass Spectrometry data for bacterial antibiotic resistance identification, demonstrating the technique’s capability for complex predictive modeling from spectral data [25]. Collectively, these studies indicate that deep learning and transfer learning techniques applied to various biological data types (including mass spectrometry, genomic, and protein sequence data) can significantly enhance the prediction of antibiotic resistance. López-Cortés et al. showed that their MSDeepAMR model, employing deep learning on raw MS data and leveraging transfer learning for adaptation to external datasets, achieves significantly enhanced prediction accuracy for antibiotic resistance. [26]. Mesureur et al. developed a comprehensive Brucella MALDI-TOF MS reference database using a specific biomathematical algorithm (ASC) to overcome intra-genus spectral similarity [8]. This approach significantly improved species-level identification accuracy for clinically relevant Brucella species and allowed effective differentiation from the closely related Ochrobactrum genus, enhancing the precision of clinical diagnostics for brucellosis. Therefore, optimizing microbiological research and clinical diagnostics can be achieved through advanced analysis of spectral data; studies demonstrate that employing machine learning techniques, potentially augmented by feature engineering (e.g., preprocessing ensembles or dynamic binning) and transfer learning, allows for the effective identification and prediction of bacterial antimicrobial resistance (AMR) directly from MALDI-TOF MS data, thereby providing more precise and rapid support for clinical decision-making. [27–29]. Motivated by these findings, this study aims to integrate MALDI-TOF MS with transfer learning to enhance the prediction of species-specific bacterial antimicrobial resistance. A key aspect of our proposed methodology involves leveraging genus-level information, hypothesizing that this approach can improve the accuracy and robustness of AMR predictions for individual bacterial species.

### Experimental Section Datasets

The data used in this study were obtained from the DRIAMS dataset, collected from four diagnostic laboratories in Switzerland between 2016 and 2018 [27], containing MALDI-TOF MS data from >300,000 clinical isolates and 768,300 AMR labels. All data used in this study were accessed between February 1, 2024 and February 29, 2024 for research purposes. This study was based on a retrospective analysis of fully anonymized medical records. The study protocol was reviewed and approved by the Research Ethics Committee of Taichung Tzu Chi Hospital, Taiwan, and was granted exempt status (IRB number: REC112-51), with the requirement for informed consent waived. The data included 803 different bacterial and fungal pathogens, which were divided into four subsets (DRIAMS-A–DRIAMS-D). The MS data were preprocessed by grouping them into intervals of 3 Daltons (Da) each, ranging from 2,000 to 20,000 Da, which resulted in a 6,000-dimensional vector representation. AMR labels were converted to binary labels (susceptible and resistant) and divided into training and testing sets. Figure 1 presents the overall workflow of the study, encompassing data collection, filtering, spectrum processing, and AMR prediction results. For each pathogen, we selected clinically relevant antibiotics for testing and created a DRIAMS subset for each antibiotic. These subsets were further divided into training and testing datasets using the methods mentioned above. For *Staphylococcus aureus*, we concentrated on three clinically implicated antibiotics: oxacillin, fusidic acid, and ciprofloxacin. For *Klebsiella pneumoniae*, we focused on two antibiotics: ciprofloxacin and ceftriaxone. A deep neural network classifier was used to predict antibiotic resistance, with hyperparameter tuning conducted to optimize the model. K-fold cross-validation was used to assess the robustness of the model, providing a more comprehensive evaluation of the model’s stability and generalizability. Performance was evaluated using the area under the receiver operating characteristic curve (AUROC) and area under the precision-recall curve (AUPRC), while Shapley values were used to assess the contribution of each feature to the AMR prediction.

**Figure 1.**
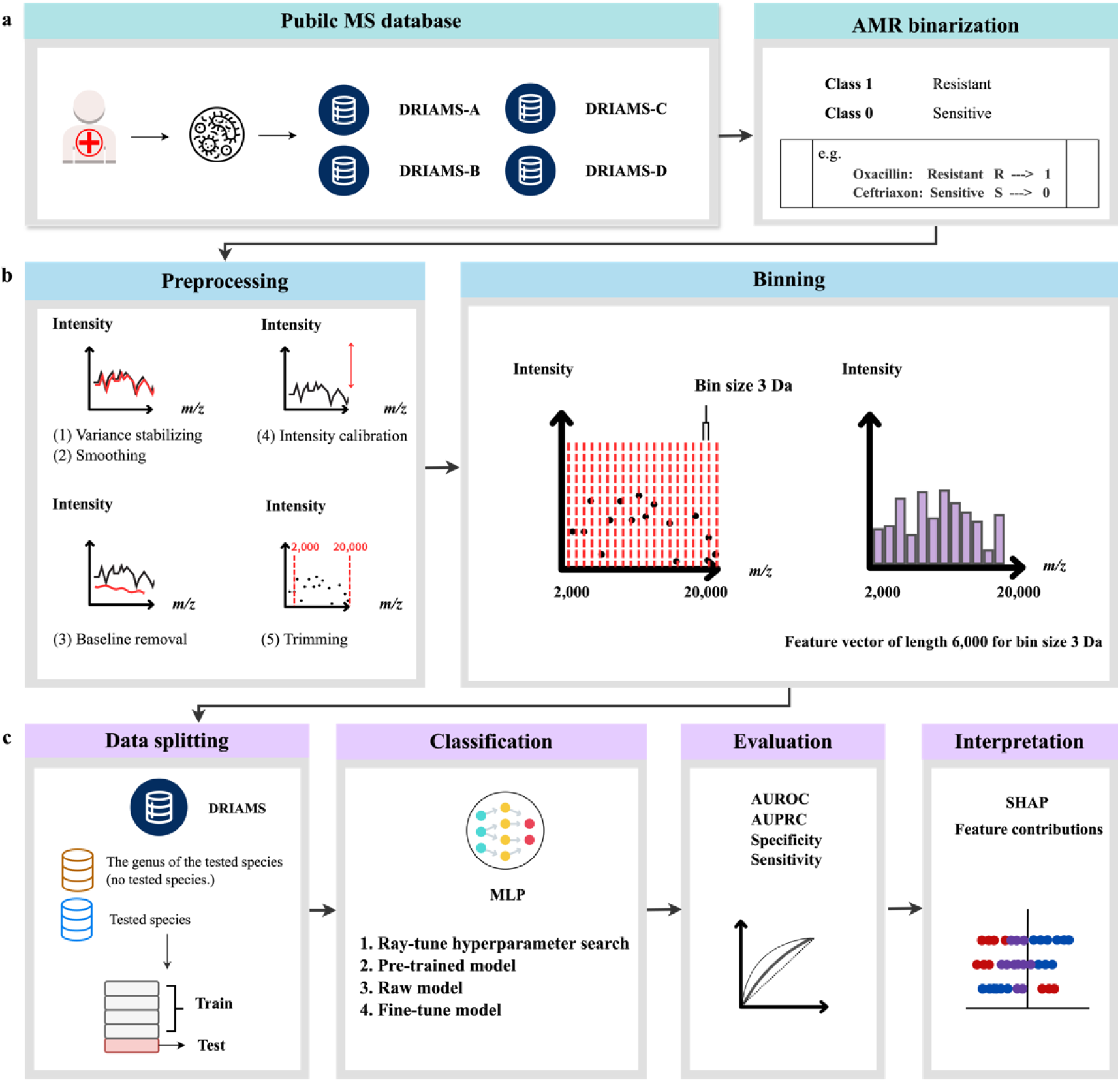
Workflow for AMR prediction using mass spectrum. **(A)** AMR Binarization: AMR is modeled as a binary classification scenario, with category 1 representing resistance and category 0 representing sensitivity. (**B)** Preprocessing: Cleaning of mass spectrometry data. Binning: Spectra are binned into equally sized feature vectors suitable for deep learning. (**C)** Data Splitting: The dataset is reduced to include samples from only two pathogens for this study. The data is divided into an 80% training set and 20% testing set. Classification: A deep neural network classifier (multilayer perceptron) was used for hyperparameter search, employing ten-fold cross-validation to train the classifier. Evaluation: Model performance is assessed using common metrics (AUROC and AUPRC). Interpretation: Shapley values are utilized to assess the contribution of individual features to AMR prediction. AMR, antimicrobial resistance; AUROC, area under the receiver operating characteristic curve, AUPRC, area under the precision-recall curve; SHAP, Shapley additive explanation.

### Model Definition

The modeling approach began with the Raw Model, a foundational model trained specifically on data from the target bacterial species to predict antibiotic resistance, providing targeted analysis and predictive capabilities tailored to the species. Subsequently, the Pretrained Model was developed using data from all bacterial genera, excluding the target species, to predict antibiotic resistance against the target antibiotic. This stage aims to capture the broader influences of other bacterial species on antibiotic resistance characteristics, enriching the model with diverse bacterial data. In the final step, the Finetuned Model utilized the training weights from the Pretrained Model, applying them to the target species’ data through transfer learning. This process leveraged knowledge gained from other bacterial species to refine the model’s predictive performance, enhancing its accuracy in predicting antibiotic resistance for the target species.

### Evaluation metrics

This study primarily employed two evaluation metrics to assess the model performance: AUROC and AUPRC [30]. The AUROC is a metric used to assess the performance of the classification model. It is derived from the relationship between the true positive rate (or sensitivity) and the false positive rate across various threshold levels. The ROC curve is a graphical representation illustrating the diagnostic ability of a binary classifier system, with true positive rate plotted on the *y*-axis and false positive rate on the *x*-axis. AUROC values range from 0.5 to 1.0: an AUROC of 0.5 indicates that the model has no discriminative ability, equivalent to random guessing, while values closer to 1.0 indicate better model performance, suggesting that the model can effectively differentiate between positive and negative classes.

The AUPRC is particularly useful for evaluating models on datasets with imbalanced classes. The precision-recall curve illustrates the tradeoff between precision—the proportion of true positive results among all positive results predicted by the model—and recall—the proportion of true positive results among all actual positives—across various threshold levels. The AUPRC values range from 0 to 1, with higher values indicating better performance. A high AUPRC suggests that the model maintains a good balance between precision and recall, making it particularly informative when dealing with data where the negative class significantly outnumbers the positive class. To evaluate the model, we introduced the following formulas:

Precision (*PRE*) is defined as:

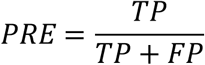

where *TP* is the true positive count and *FP* represents the false positive count. This metric indicates the ratio of correctly identified positive samples to the total number of recommendations.

Recall (*REC*) is defined as:

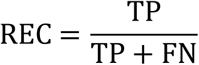

where *FN* is the false negative count. This metric indicates the ratio of correctly identified positive samples to the total number of positive samples.

F1 Score (*F*_1_) is the harmonic mean of precision and recall and is defined as

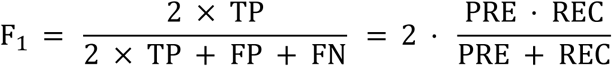

Precision and recall should ideally be close to one. However, in practice, achieving high precision typically results in lower recall and vice versa.

### Interpretability with Shapley values

The Shapley additive explanations (SHAP) algorithm was employed to interpret the data, allowing us to obtain Shapley values for each sample and enhance the interpretability of the classification models. These Shapley values demonstrate the influence of individual features on the overall output of the model. Based on cooperative game theory, Shapley values have substantial applications not only in the distribution of cooperative gains, but also in calculating the weighted average of the marginal contributions of each feature across all possible subsets. This is considerably important for further research and practical applications. Ayad et al. investigated the use of Shapley values in multioutput classification tasks, highlighting the significance of including label interdependencies [31]. Similarly, Michiels et al. examined the relationships between models and features, proposing a novel algorithm to enhance the explanatory power of Shapley values [32]. Another relevant study by Madakkatel and Hyppönen introduced a feature selection method based on Shapley values to identify informative features and minimize noise [33]. The *i*-th Shapley value was expressed as follows:

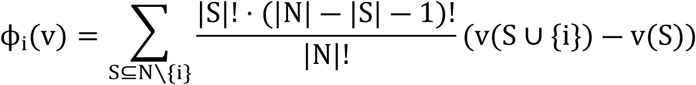

where *N* is the set of players, *v* is the characteristic function that defines the value of each subset, S ⊆ ∖ {i} denotes all subsets excluding player i, |*S*| is the size of subset S, |*N*| is the total number of players, v(*S* ∪ *i*) is the value when player i joins subset S, and v(*S*) is the value of subset S.

After completing model training, we conducted model interpretation by extracting SHAP values specifically for the class representing antibiotic susceptibility. This process involved ensuring that all input data were consistently scaled to maintain the model’s stability and reliability, preventing bias from data of varying magnitudes. Additionally, we identified the 30 most important features from the selected models, highlighting their contributions to the predictions. This comprehensive approach improved the accuracy of data interpretation and reinforced the reliability of the results, ensuring that the analysis accurately represented the influence of key features on predicting antibiotic resistance [34].

### Experimental combinations

Various bacteria-antibiotic combinations of clinical significance were investigated to evaluate the corresponding antimicrobial effects and resistance. Species within the same genus often exhibit similar characteristics, therefore, genus-level information was incorporated into the training model to enhance the robustness and accuracy of AMR prediction. This allowed for the learning of broader resistance patterns, along with the application of this information for the effective prediction of resistance. The selected bacteria included *S. aureus*, *Escherichia coli*, and *K. pneumoniae*, which are listed as priority pathogens by the World Health Organization. These were analyzed for their sensitivity and resistance to multiple antibiotics. Due to the limited sample size, five combinations of antibiotics and bacteria were selected for detailed analysis (Table 1). This approach was used to leverage genus-specific shared traits to improve predictive performance, even with limited data.

**Table 1.**
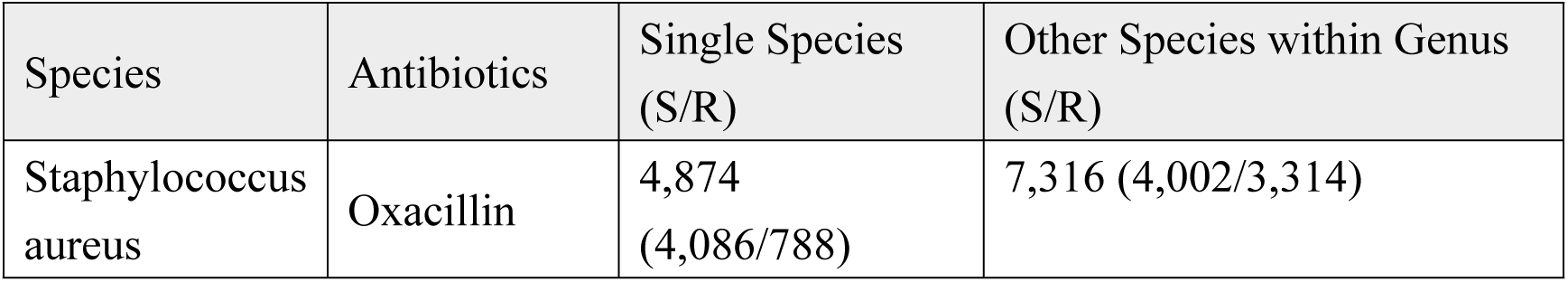
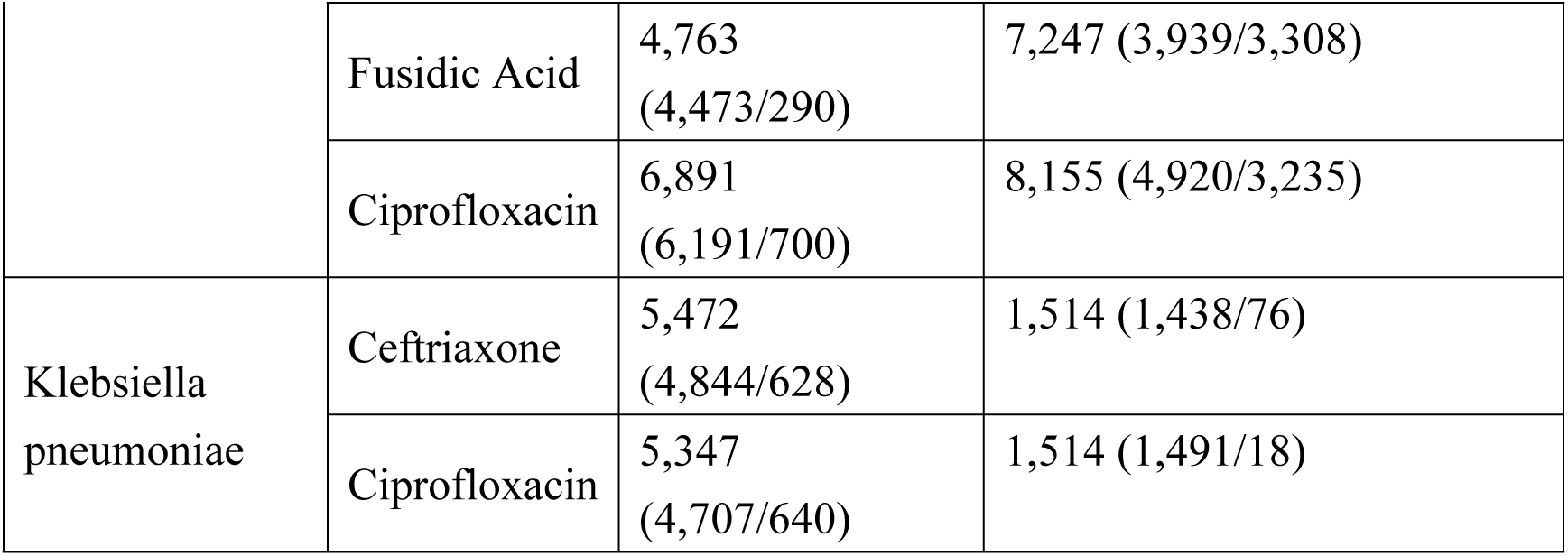
Statistical analysis of bacterial strains and antibiotics across five experimental groups Abbreviations: S = Sensitive, R = Resistant.

The analysis results showed that the number of *S. aureus* strains sensitive to oxacillin, fusidic acid, and ciprofloxacin was 4,086, 4,473, and 6,191, respectively, while the number of resistant strains was 788, 290, and 700, respectively. Owing to the insufficient sample size of *E. coli*, further analyses were not conducted (Supplementary Table S1). For *K. pneumoniae*, the numbers of strains sensitive to ceftriaxone, cefepime, tobramycin, ciprofloxacin, and meropenem were 4,844, 4,785, 2,527, 4,707, and 3,281, respectively, whereas the numbers of resistant strains were 628, 243, 317, 640, and 24, respectively (Table 1). Three antibiotic combinations were ultimately selected for *S. aureus*, and two were selected for *K. pneumoniae* for in-depth quantitative analysis.

### Statistical methods

Five- and ten-fold cross-validations were employed to calculate the mean performance and its standard deviation in the test set. Cross-validation is a technique that divides a dataset into multiple subsamples; most samples are used to train the model, whereas the remaining samples are used to validate the model [35]. This method can effectively reduce model overfitting and ensure the reliability and stability of evaluation results [36,37]. Table 2 shows the results of the five-fold cross-validation. Ten-fold cross validation reveal similar results in Supplementary Table S6.

**Table 2.**
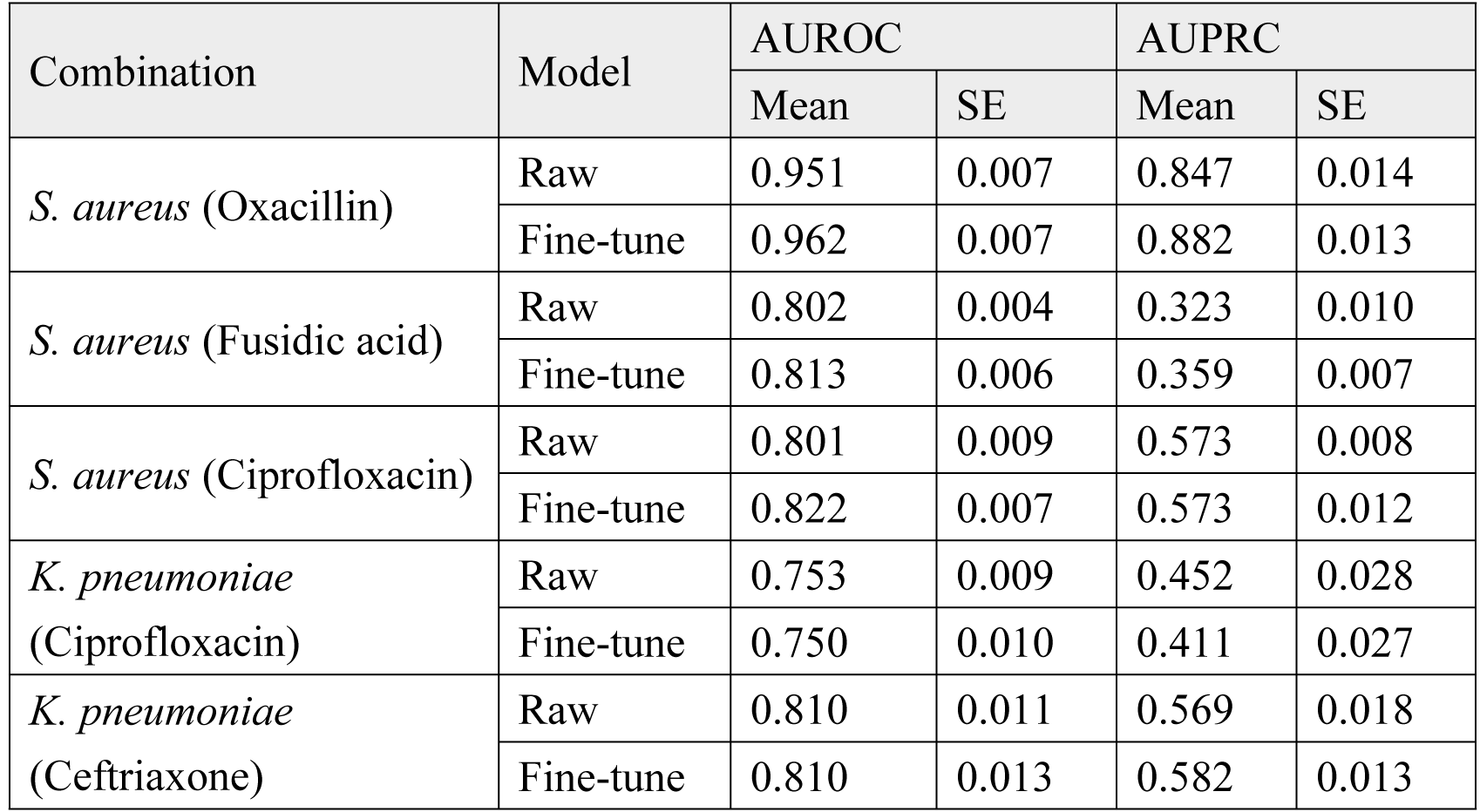
Comparison of Performance Metrics: Raw vs. Fine-tuned Models under five-fold cross-validation Abbreviations: *S. aureus = Staphylococcus aureus, K. pneumoniae* = Klebsiella pneumoniae, SE, standard error, AUROC = area under the receiver operating characteristic curve, AUPRC = area under the precision-recall curve, Raw: A model that does not use the training weights of a pre-trained model, Fine-tune: A model that uses the training weights of a pre-trained model.

### Deep learning methods

A multilayer perceptron deep neural network (DNN) was used to train the DRIAMS dataset for the binary classification study of bacterial resistance. This approach focused on the MALDI-TOF MS data of bacteria belonging to the same genus to enhance antibiotic resistance prediction at the species level by leveraging the genus-specific shared characteristics and resistance patterns.

In the early stages of model training, pretraining was conducted using all species within the genus of the test species (excluding the test species itself) from the training dataset to build an initial resistance prediction model. This pretraining phase aimed to capture broader resistance characteristics. Subsequently, the weights from the pretrained models were used for fine-tuning to further train the model for the test species. Finally, the performances of models trained solely using the target species dataset and those fine-tuned using genus-level pretrained models were compared to evaluate the effect of fine-tuning on prediction accuracy.

In the process of training, RayTune, a Python package designed for scalable hyperparameter tuning, was utilized to systematically search for optimal parameter combinations [38]. This tool efficiently handles large-scale hyperparameter tuning, significantly improving the efficiency and effectiveness of model training. Finally, cross-validation techniques were used to mitigate the risk of overfitting. By repeatedly partitioning the training and validation data, these techniques ensured that the model exhibited good generalizability and robustness [39]. The comprehensive application of these techniques effectively enhanced the accuracy and stability of the model (Supplementary Table S2).

### Addressing data imbalance

We addressed the issue of data imbalance in this study, using the adaptive synthetic sampling (ADASYN) technique to enhance the quantity and diversity of minority class samples. Data imbalance is a common challenge in machine learning, particularly in classification tasks, as it often leads to a model bias towards the majority class, thereby reducing the prediction accuracy for the minority class [38]. The core of the ADASYN technique functions by dynamically generating various numbers of new samples based on the computation of the K-nearest neighbors of each minority class sample, a computation that utilizes neighbor distances. This approach increases the number of minority class samples and ensures the diversity and authenticity of the dataset. The ADASYN technique was specifically applied to our dataset through the following steps: (1) a K value, typically 5 or 10, was selected; (2) the K-nearest neighbors for each minority class sample were calculated, and (3) new samples were generated dynamically based on the distances. The results of the study showed that the application of the ADASYN technique led to a more balanced class distribution in the dataset, and the model exhibited significant improvements in key metrics, such as accuracy, AUROC, and AUPRC [39]. These demonstrate the effectiveness of ADASYN in addressing data imbalance, thereby enhancing model stability and prediction accuracy thus highlighting its value for related future research [40].

## Results

Based on the data presented in Table 1, the sensitivity and resistance (S/R) ratios for *S. aureus* are as follows: 4,086/788 for oxacillin, 4,473/290 for fusidic acid, and 6,191/700 for ciprofloxacin. For *K. pneumoniae,* the S/R ratios are 4,844/628 for ceftriaxone and 4,707/640 for ciprofloxacin. These distributions suggest a notable imbalance in the data.

Figure 1 illustrates the workflow for AMR prediction using mass spectrometry. AMR was binarized into resistant (category 1) and sensitive (category 0). The MS data was then cleaned and binned into 6000 feature vectors. The dataset was split into 80% training set and 20% testing set.

Table 2 presents the comparison between raw and fine-tune models for different bacteria in predicting the antibiotics resistance with K-fold cross-validation. In *S. aureus*, the fine-tuned model achieved an AUROC of 0.921 for predicting resistance to oxacillin, compared to 0.919 with the raw model under K=5. Similarly, with K=10, the fine-tuned model reached an AUROC of 0.928, slightly outperforming the raw model’s 0.926 (as shown in Supplementary Table S6). For fusidic acid, although the raw model’s AUROC was 0.811 in 10-fold, the fine-tuned model slightly improved to 0.822, indicating that fine-tuning positively improved model performance. In the evaluation of ciprofloxacin, the fine-tuned model enhance model performance, with AUROCs of 0.822 and 0.832 in 5-fold and 10-fold, respectively, compared to those of the raw model AUROCs of 0.801 in 5-fold and 0.810 in 10-fold. For *K. pneumoniae*, the fine-tuned model for ceftriaxone yielded AUPRCs of 0.582 in 5-fold and 0.596 in 10-fold, indicating better performance in the prediction. Overall, the experimental results showed that fine-tuned models with genus level data could enhance prediction performance (Fig. 2).

**Figure 2.**
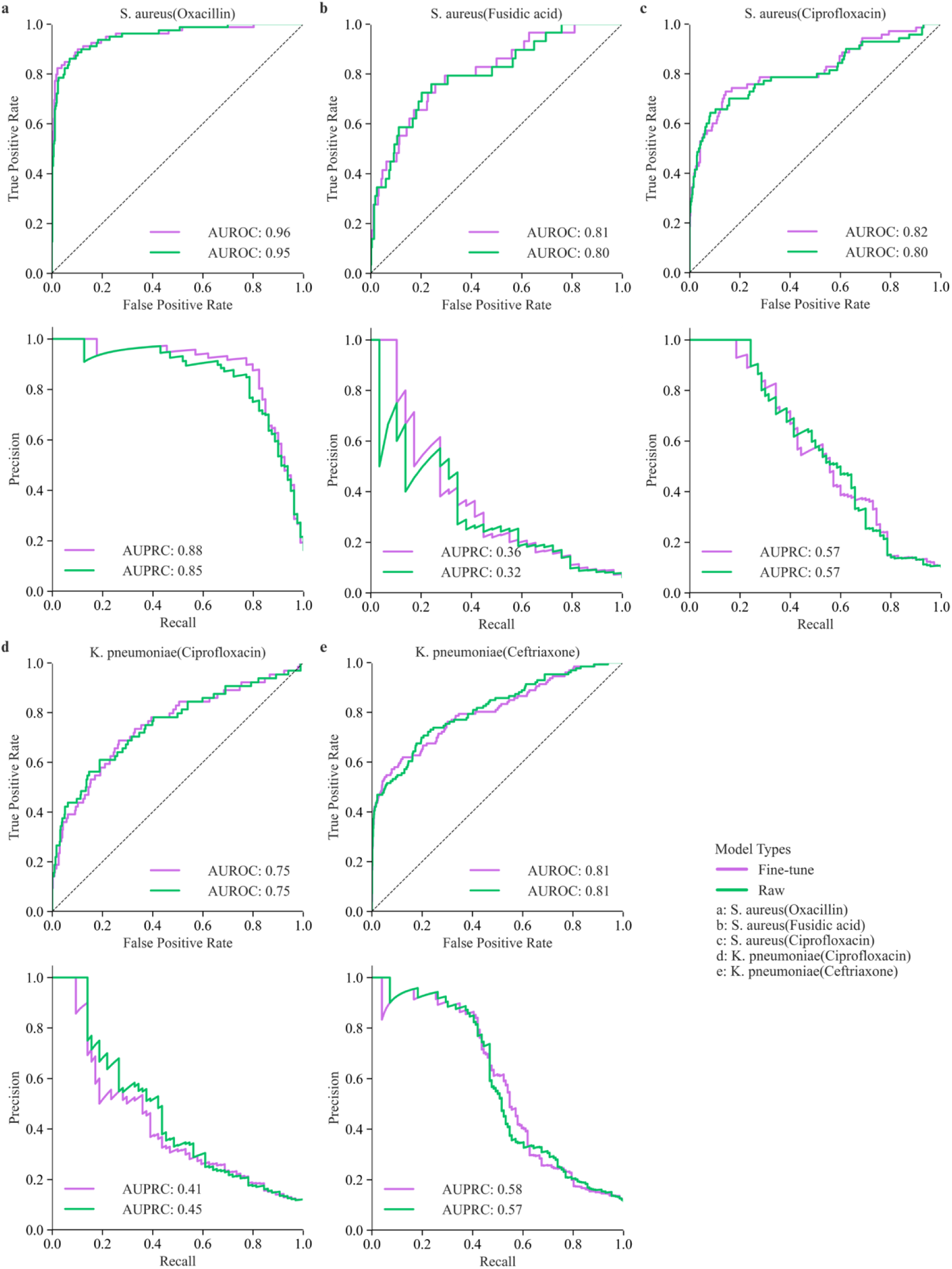
Receiver operating characteristic and precision-recall curves for fine-tuned and raw models. **(A)** Comparing AUROC and AUPRC for *S. aureus* resistance to oxacillin, the fine-tuned model reached an AUROC of 0.96 and an AUPRC of 0.88, whereas the raw model had an AUROC of 0.95 and an AUPRC of 0.85. Hence, the fine-tuned model demonstrates superior performance. (**B)** For *S. aureus* resistance to fusidic acid, the fine-tuned model exhibited an AUROC of 0.81 and an AUPRC of 0.36, whereas the raw model had an AUROC of 0.80 and an AUPRC of 0.32. The fine-tuned model demonstrates superior performance. (**C)** For *S. aureus* resistance to ciprofloxacin, the fine-tuned model had an AUROC of 0.82 and an AUPRC of 0.57, whereas the raw model had an AUROC of 0.80 and an AUPRC of 0.57, showing a marginal improvement in AUROC for the fine-tuned model. (**D)** For the resistance of *K. pneumoniae* to ciprofloxacin, the fine-tuned model reached an AUROC of 0.75 and an AUPRC of 0.41, compared to 0.75 and 0.45, respectively, for the raw model. The fine-tuned model does not show superior performance. (**E)** For *K. pneumoniae* resistance to ceftriaxone, the two models showed identical AUROCs of 0.81; however, the fine-tuned model had a slightly higher AUPRC of 0.58 compared to 0.57 for the raw model, indicating no significant improvement with the fine-tuned model. *S. aureus*, *Staphylococcus aureus*; *K. pneumoniae*, *Klebsiella pneumoniae*; AUROC, area under the receiver operating characteristic curve; AUPRC, area under the precision-recall curve.

ADASYN improved the predictive performance of most antibiotics as demonstrated in Supplementary Table S3. For *Staphylococcus aureus*, the use of oxacillin increased the AUROC from 0.94 to 0.95, while the AUPRC remained unchanged at 0.85. For fusidic acid, the AUROC increased from 0.70 to 0.80, and the AUPRC increased from 0.18 to 0.32. In the assessment of ciprofloxacin, both AUROC and AUPRC slightly increased to 0.80 and 0.57, respectively. For *K. pneumoniae*, both AUROC and AUPRC for ciprofloxacin also showed a slight improvement.

The SHAP value analysis revealed differences between the fine-tuned models and raw models in the top-30 important-feature selections, indicating that the fine-tuned models prioritized specific features to enhance prediction performance. The overlap of features was 90% for oxacillin, 50% for fusidic acid, and 56.67% for ciprofloxacin in *S. aureus*. For ciprofloxacin in *K. pneumoniae*, the overlap was 70%, and 46.67% for ceftriaxone (Supplementary Table S4). These differences were also evident in the similarities observed among the top five MS data features. Using oxacillin for *S. aureus*, the top five overlapping mass spectral feature ranges were 2,135–2,138 m/z, 3,212–3,215 m/z, 6,893–6,896 m/z, and 2,243–2,246 m/z; using fusidic acid, the mass ranges were 5,504–5,507 m/z and 6,896–6,899 m/z; and using ciprofloxacin, the mass ranges were 4,304–4,307 m/z, 5,528–5,531 m/z, and 4,298– 4,301 m/z. Similarly, for using ciprofloxacin *K. pneumoniae*, the top five similar mass ranges were 2,414–2,417 m/z, 2,591–2,594 m/z, 2,612–2,615 m/z, 2,135–2,138 m/z, and 3,335–3,338 m/z; and using ceftriaxone, the ranges were 2,135–2,138 m/z, 2,066– 2,069 m/z, and 2,069–2,072 m/z. These data highlight the preferences of different models for specific features, underscoring the importance of feature engineering and model tuning. These findings have important implications for understanding feature engineering and model adjustment, aiding researchers in identifying influential features under various conditions to develop more effective predictive models (Fig. 3–7).

**Figure 3.**
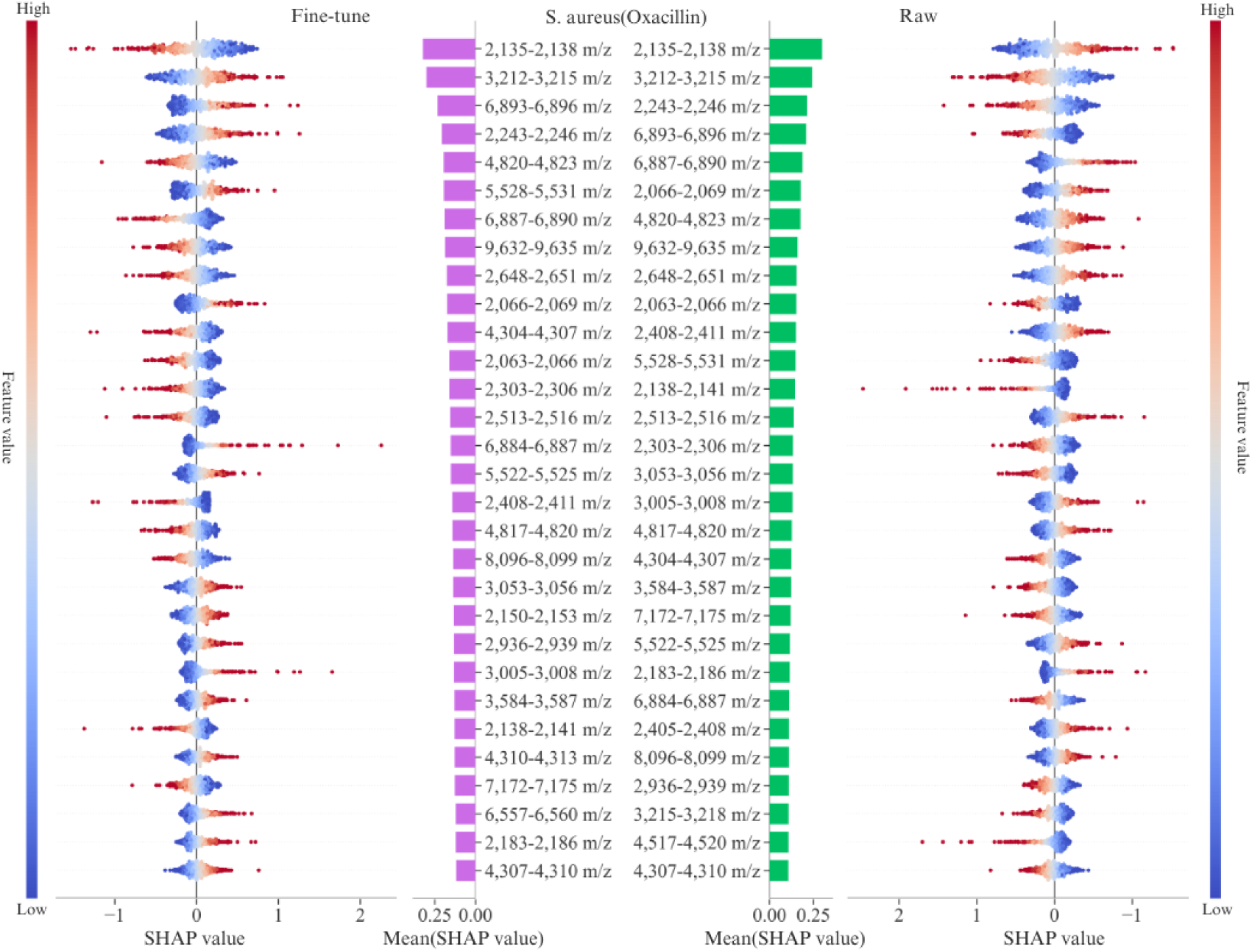
SHAP values for *S. aureus* (oxacillin). The figure shows the SHAP values for the top 30 features predicting oxacillin resistance in *S. aureus*. The left plot shows SHAP values from the fine-tuned model, whereas the right plot shows the raw SHAP values. Colors represent feature values, with red indicating high values and blue indicating low values. The central bar chart depicts the mean SHAP value importance for each m/z feature, ranked in order. The overlap of features for oxacillin is 90%. The top five overlapping feature mass ranges are 2,135–2,138 m/z, 3,212–3,215 m/z, 6,893–6,896 m/z, and 2,243–2,246 m/z. SHAP, Shapley additive explanation; *S. aureus*, *Staphylococcus aureus*.

**Figure 4.**
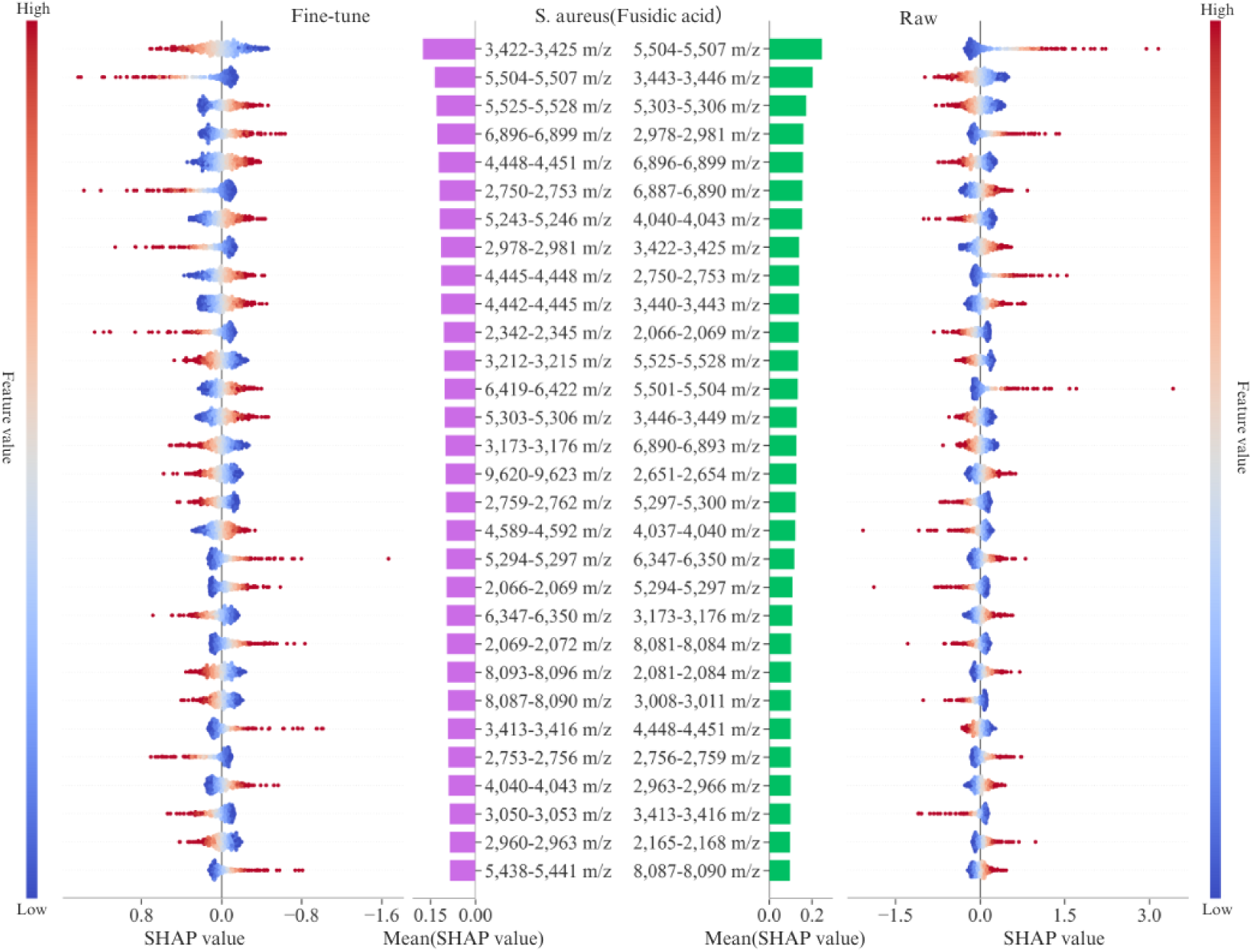
SHAP values for *S. aureus* (fusidic acid). The figure shows the SHAP values for predicting the top 30 features of fusidic acid resistance in *S. aureus*. The left plot shows the fine-tuned SHAP values, whereas the right plot shows those from raw models. Colors indicate feature values, with red denoting high values and blue denoting low values. The central bar chart indicates the importance of the mean SHAP value for each m/z feature in order. The overlap of features for fusidic acid is 50%. The top five overlapping feature mass ranges are 5,504–5,507 m/z and 6,896– 6,899 m/z. SHAP, Shapley additive explanation; *S. aureus*, *Staphylococcus aureus*.

**Figure 5.**
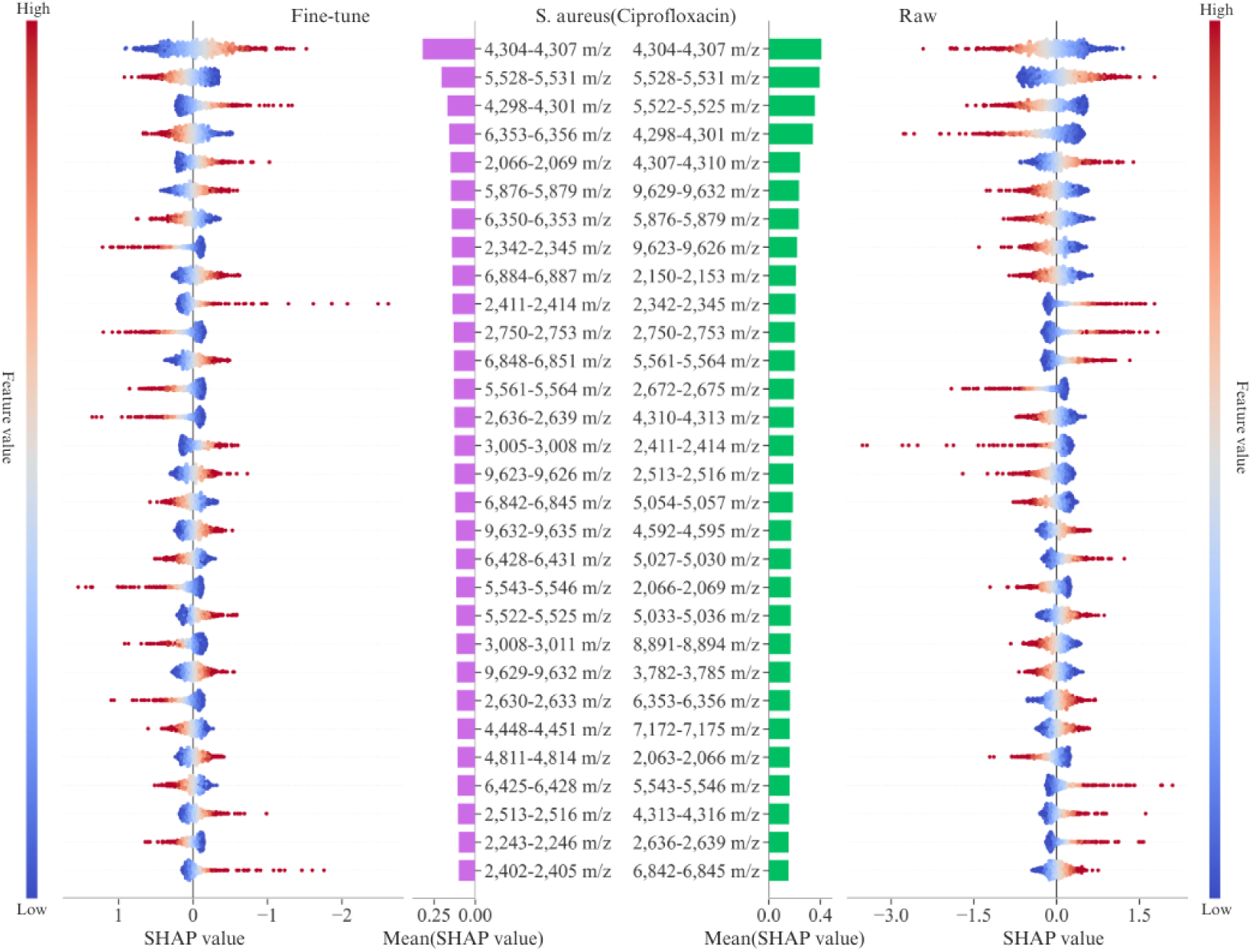
SHAP values for *S. aureus* (ciprofloxacin). The figure shows the SHAP values for predicting the top 30 features associated with ciprofloxacin resistance in *S. aureus*. The left plot shows the SHAP values for the fine-tuned model, whereas the right plot shows the raw SHAP values. Colors represent feature values, with red indicating high values and blue indicating low values. The central bar chart shows the mean SHAP value importance for each m/z feature in order. The overlap of ciprofloxacin features in *S. aureus* is 56.67%. The top five overlapping feature mass ranges include 4,304–4,307 m/z, 5,528–5,531 m/z, and 4,298–4,301 m/z. SHAP, Shapley additive explanation; *S. aureus*, *Staphylococcus aureus*.

**Figure 6.**
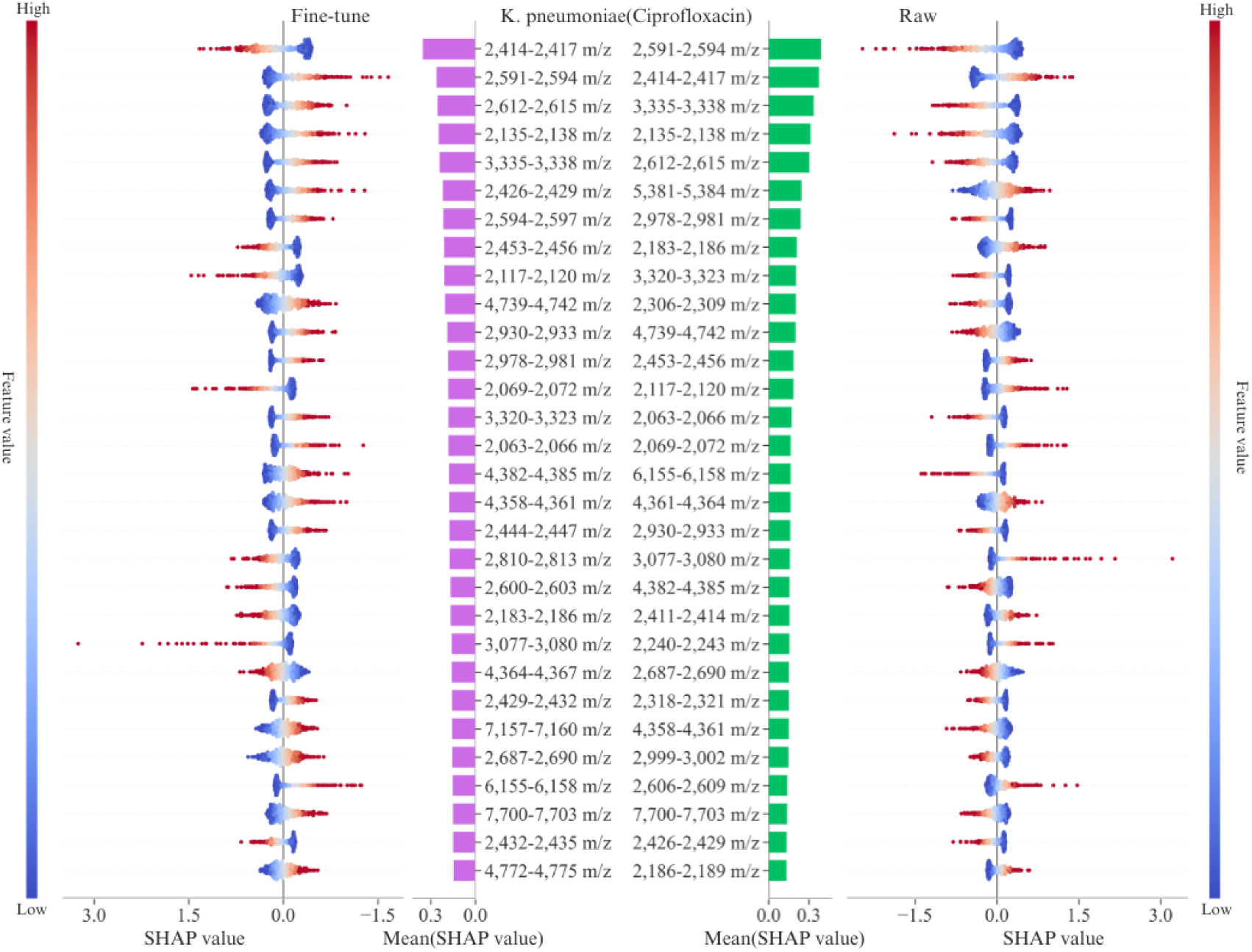
SHAP values for *K. pneumoniae* (ciprofloxacin). The figure shows the SHAP values for predicting the top 30 features associated with ciprofloxacin resistance in *K. pneumoniae*. The left plot shows the SHAP values for the fine-tuned model, whereas the right plot shows the raw SHAP values. Colors represent feature values, with red indicating high values and blue indicating low values. The central bar chart shows the mean SHAP value importance for each m/z feature in order. The overlap of ciprofloxacin features in *K. pneumoniae* is 70%. The top five overlapping feature mass ranges include 2,414–2,417 m/z, 2,591–2,594 m/z, 2,612–2,615 m/z, 2,135–2,138 m/z, and 3,335–3,338 m/z. SHAP, Shapley additive explanation; *K. pneumoniae*, *Klebsiella pneumoniae*.

**Figure 7.**
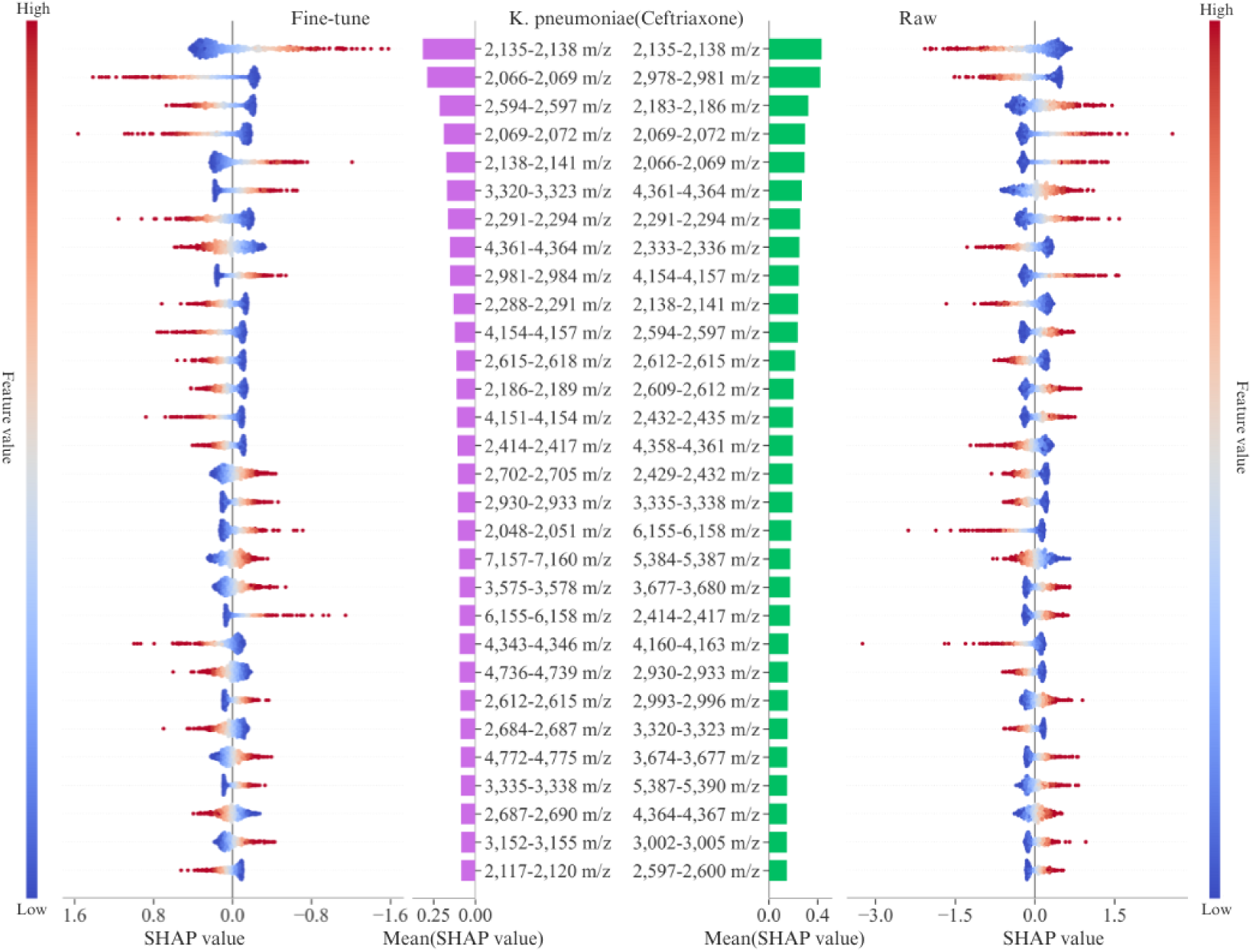
SHAP values for *K. pneumoniae* (ceftriaxone). The figure shows the SHAP values for predicting the top 30 features associated with ceftriaxone resistance in *K. pneumoniae*. The left plot shows the SHAP values for the fine-tuned model, whereas the right plot shows the raw SHAP values. Colors represent feature values, with red indicating high values and blue indicating low values. The central bar chart shows the mean SHAP value importance for each m/z feature in order. The overlap of ceftriaxone features in *K. pneumoniae* is 46.67%. The top five overlapping feature mass ranges include 2,135–2,138 m/z, 2,066–2,069 m/z, and 2,069–2,072 m/z. SHAP, Shapley additive explanation; *K. pneumoniae*, *Klebsiella pneumoniae*.

## Discussion

We explored the potential of DNNs for predicting antibiotic resistance in bacterial species by combining genus-level microbiological data with transfer learning techniques. The model was pretrained for broader genus-specific resistance characteristics using the data from other species within the same genus. Subsequently, species-specific training was performed using the pretrained model to enhance the antibiotic resistance prediction. The fine-tuned model with genus data exhibited notably enhanced prediction accuracy for *S. aureus* for various antibiotics compared to the raw model trained only on *S. aureus* data. The AUROC values for predicting resistance to oxacillin, fusidic acid, and ciprofloxacin were notably high, with the fine-tuned model outperforming the raw model in each case. Specifically, the AUROC for oxacillin was 0.962 versus 0.951 for the raw model, 0.813 versus 0.802 for fusidic acid, and 0.822 versus 0.801 for ciprofloxacin. Moreover, the AUPRC for certain antibiotic combinations demonstrated significant improvements. For instance, the fine-tuned model consistently outperformed the raw model in all three selected antibiotics for *S. aureus*. Similarly, for *K. pneumoniae*, the fine-tuned model achieved an AUPRC of 0.582 for ceftriaxone, compared to 0.569 for the raw model. These results highlight the effectiveness of incorporating transfer learning to leverage intra-genus data and provide plausible microbiological insights into genus-specific shared resistance mechanisms.

Overall, our method performed excellently in most antibiotic resistance predictions, displaying substantial advantages, particularly for certain antibiotics, such as oxacillin and fusidic acid. Similarly, the approach utilized by López-Cortés et al., which incorporates DNNs and transfer learning from the DRIAMS-A subset, has shown enhanced performance when applied to subsets B and C. Specifically, for oxacillin resistance in *S. aureus*, the AUROC improved from 0.683 to 0.793 in subset B, and from 0.654 to 0.782 in subset C. The method employed by Caroline Weis et al. performed suboptimally in several evaluations [27]. For instance, when models trained on subset A were used to predict outcomes on subset B, the AUROC values were only 0.64 and 0.68, indicating relatively poor predictive performance. The comprehensive comparison among the three studies is presented in Supplementary Table S5. These findings demonstrate that the integration of deep learning with transfer learning provides high accuracy and reliability in predicting antibiotic resistance, particularly when fine-tuned with genus-level data. Nonetheless, future research should further investigate the applicability of this approach across diverse datasets to assess its potential for enhancing the accuracy of AMR predictions.

Future investigation should also focus on using MALDI-TOF MS with expanded databases and broader genus-level data to improve species-level predictions. Herein, the clinically significant genus *Staphylococcus* was investigated, and the data from multiple species within this genus were used to improve the accuracy of antibiotic resistance prediction, addressing data limitations and highlighting genus-specific shared resistance traits. Therefore, incorporating more pathogen-antibiotic combinations, such as those used in this study, may provide a more comprehensive evaluation of the effects of transfer learning techniques on prediction accuracy.

Additionally, integrating multimodal data, including genomic sequencing and clinical data, may further enhance prediction accuracy and provide deeper insights into resistance mechanisms. Overall, based on the results of this study, we propose employing transfer learning and fine-tuning for augmenting sparse data. These improved approaches may predict antibiotic resistance with greater accuracy to better support clinical diagnosis and treatment, thereby aiding in the effective management of infections with AMR pathogens.

This study has some limitations. First, the scope was mainly confined to two bacterial species, *S. aureus* and *K. pneumoniae*, as well as a selected set of antibiotics, which may not have fully captured the diversity of pathogens and antibiotic resistance mechanisms encountered in clinical settings. Second, the DRIAMS dataset used may have introduced geographical and temporal biases, limiting the generalizability of the model for other regions and bacterial strains. Third, some antibiotic-pathogen combinations had limited sample sizes, particularly for antibiotic-resistant strains, which could have affected the performance and reliability of their corresponding predictions. Finally, although the models demonstrated effective predictive ability, insights into the underlying mechanisms of antibiotic resistance remain elusive. The integration of other omics data and biological information may enhance the interpretability and actionability of the results.

## Conclusion

Altogether, the findings of this study showed the potential of combining deep learning and transfer learning techniques in predicting antibiotic resistance using MALDI-TOF MS data. Furthermore, they highlighted the effectiveness of incorporating the genus-level data to enhance species-level predictions, which addressed the challenge of limited species-specific data by leveraging genus-specific broader characteristics. Future research should focus on applying this approach to include a wider range of pathogens and antibiotics. Further refining transfer learning techniques should also be undertaken, and additional data types should be integrated. Implementing these steps would facilitate the development of predictive models with enhanced accuracy and applicability in clinical settings. Addressing these aspects of research is crucial for advancing the field and for achieving clinical utility of MALDI-TOF MS technology for the rapid prediction of antibiotic resistance.

## Data Availability

The data underlying the results presented in the study are available from https://doi.org/10.5061/dryad.bzkh1899q

## Acknowledgments Data statement

All data can be accessed and downloaded via the following link: https://doi.org/10.5061/dryad.bzkh1899q.

## Author contributions

## References

1. Zhu Y, Huang WE, Yang Q. Clinical perspective of antimicrobial resistance in bacteria. Infect Drug Resist. 2022;15:735–746. 10.2147/IDR.S345574.

2. Kongnakorn T, Tichy E, Kengkla K, et al. Economic burden of antimicrobial resistance and inappropriate empiric treatment in Thailand. Antimicrob Steward Healthc Epidemiol. 2023;3:e109. 10.1017/ash.2023.169.

3. Llor C, Bjerrum L. Antimicrobial resistance: risk associated with antibiotic overuse and initiatives to reduce the problem. Ther Adv Drug Saf. 2014;5:229–241. 10.1177/2042098614554919.

4. Spellberg B, Bartlett JG, Gilbert DN. The future of antibiotics and resistance. N Engl J Med. 2013;368:299–302. 10.1056/NEJMp1215093.

5. Karkman A, Pärnänen K, Larsson DGJ. Fecal pollution can explain antibiotic resistance gene abundances in anthropogenically impacted environments. Nat Commun. 2019;10:80. 10.1038/s41467-018-07992-3.

6. World Health Organization (WHO). Antimicrobial resistance. Accessed 2023. https://www.who.int/news-room/fact-sheets/detail/antimicrobial-resistance.

7. Paul S, Singh P, Sharma S, et al. MALDI-TOF MS-based identification of melanized fungi is faster and reliable after the expansion of in-house database. Proteomics Clin Appl. 2019;13:e1800070. 10.1002/prca.201800070.

8. Mesureur J, Arend S, Cellière B, et al. A MALDI-TOF MS database with broad genus coverage for species-level identification of Brucella. PLOS Negl Trop Dis. 2018;12:e0006874. 10.1371/journal.pntd.0006874.

9. Calderaro A, Chezzi C. MALDI-TOF MS: a reliable tool in the real life of the clinical microbiology laboratory. Microorganisms 2024;12:322. 10.3390/microorganisms12020322.

10. Iles RK, Iles JK, Zmuidinaite R. Development of a MALDI-TOF mass spectrometry test for viruses. In: Shah HN, Gharbia SE, Shah AJ, Tranfield EY, Thompson KC, eds. Microbiological identification using MALDI-TOF and tandem mass spectrometry: industrial and environmental applications. John Wiley & Sons Ltd; 2023:117–145. 10.1002/9781119814085.ch5.

11. Haider A, Ringer M, Kotroczó Z, Mohácsi-Farkas C, Kocsis T. The current level of MALDI-TOF MS applications in the detection of microorganisms: a short review of benefits and limitations. Microbiol Res. 2023:14:80–90. 10.3390/microbiolres14010008.

12. Piga I, Magni F, Smith A. The journey towards clinical adoption of MALDI-MS-based imaging proteomics: from current challenges to future expectations. FEBS lett. 2024;598:621–634. 10.1002/1873-3468.14795.

13. Nakayama K, Li X, Shimizu K, et al. qShot MALDI analysis: A rapid, simple, convenient, and reliable quantitative phospholipidomics approach using MALDI-TOF/MS. Talanta 2023;254:124099. 10.1016/j.talanta.2022.124099.

14. Kopcakova A, Stramova Z, Kvasnova S, Godany A, Perhacova Z, Pristas P. Need for database extension for reliable identification of bacteria from extreme environments using MALDI TOF mass spectrometry. Chem Pap. 2014;68:1435–42. 10.2478/s11696-014-0612-0.

15. Rychert J. Benefits and limitations of MALDI-TOF mass spectrometry for the identification of microorganisms. J Infectiology. 2019;2:1–5. 10.29245/2689-9981/2019/4.1142.

16. Raghu M, Zhang C, Kleinberg J, Bengio S. Transfusion: understanding transfer learning for medical imaging. Adv Neural Inf Process Syst. 2019;32. 10.48550/arXiv.1902.07208.

17. Mustafa B, Loh A, Freyberg J, et al. 2021. Supervised transfer learning at scale for medical imaging. arXiv preprint. 10.48550/arXiv.2101.05913.

18. Wadhwa D, Kumar S, Rengarajan A. Leveraging transfer learning for improved medical image classification. In: 2024 International Conference on Optimization Computing and Wireless Communication (ICOCWC). IEEE Publications 2024: 1–7. 10.1109/ICOCWC60930.2024.10470652.

19. Seddiki K, Saudemont P, Precioso F, et al. Towards CNN representations for small mass spectrometry data classification: from transfer learning to cumulative learning. bioRxiv 2020. 10.1101/2020.03.24.005975.

20. Akdemir A, Shibuya T; 2020. Transfer learning for biomedical question answering. In: Clef (Working Notes), Shin HC, Roth HR, Gao M, Lu L, Xu Z, Nogues I et al., eds. Deep Convolutional Neural Networks for Computer-Aided Detection: CNN Architectures, Dataset Characteristics and Transfer Learning. IEEE Transactions on Medical Imaging, 2016;35: 1285–1298.

21. Ebbehoj A, Thunbo MØ, Andersen OE, Glindtvad MV, Hulman A. Transfer learning for non-image data in clinical research: a scoping review. PLOS Digit Health. 2022;1:e0000014. 10.1371/journal.pdig.0000014.

22. Hamid MN, Friedberg I. Transfer learning improves antibiotic resistance class prediction. Biorxiv. 2020:2020–2004. 10.1101/2020.04.17.047316.

23. Wang HY, Hsieh TT, Chung CR, et al. Efficiently predicting vancomycin resistance of *Enterococcus faecium* from MALDI-TOF MS spectra using a deep learning-based approach. Front Microbiol. 2022;13:821233. 10.3389/fmicb.2022.821233.

24. Ren Y, Chakraborty T, Doijad S, et al. Deep transfer learning enables robust prediction of antimicrobial resistance for novel antibiotics. Antibiotics (Basel). 2022;11:1611. 10.3390/antibiotics11111611.

25. López-Cortés XA, Manríquez-Troncoso JM. Deep learning for the identification of multidrug resistance in MALDI-TOF MS samples of *Escherichia coli*. 10.52591/lxai202312102.

26. López-Cortés XA, Manríquez-Troncoso JM, Hernández-García R, Peralta D. MSDeepAMR: antimicrobial resistance prediction based on deep neural networks and transfer learning. Front Microbiol. 2024;15:1361795. 10.3389/fmicb.2024.1361795.

27. Weis C, Cuénod A, Rieck B, et al. Direct antimicrobial resistance prediction from clinical MALDI-TOF mass spectra using machine learning. Nat Med. 2022;28:164–174. 10.1038/s41591-021-01619-9.

28. Chung CR, Wang HY, Chou PH, et al. Towards accurate identification of antibiotic-resistant pathogens through the ensemble of multiple preprocessing methods based on MALDI-TOF spectra. Int J Mol Sci. 2023;24:998. 10.3390/ijms24020998.

29. Nguyen HA, Peleg AY, Song J, et al. Predicting *Pseudomonas aeruginosa* drug resistance using artificial intelligence and clinical MALDI–TOF mass spectra. bioRxiv. 2023:2023–10.

30. Fawcett T. An introduction to ROC analysis. Pattern Recognit Lett. 2006;27:861–174. 10.1016/j.patrec.2005.10.010.

31. Ayad CW, Bonnier T, Bosch B, Read J. Shapley chains: extending Shapley values to classifier chains. In: International Conference on Discovery Science. Cham: Springer Nature Switzerland; 2022:541–55.

32. Michiels J, Suykens J, De Vos M. Explaining the model and feature dependencies by decomposition of the Shapley value. Decis Support Syst. 2024;182:114234. 10.1016/j.dss.2024.114234.

33. Madakkatel I, Hyppönen E. LLpowershap: logistic loss-based automated Shapley values feature selection method. arXiv preprint. 2024. 10.48550/arXiv.2401.12683.

34. Lundberg SM, Lee SI. A unified approach to interpreting model predictions. Adv Neural Inf Process Syst. 2017;30.

35. Ghojogh B, Crowley M. The theory behind overfitting, cross validation, regularization, bagging, and boosting: tutorial. arXiv preprint. 2019. 10.48550/arXiv.1905.12787.

36. Prakash K, Kalaiarasan C. Web services performance prediction with confusion matrix and K-fold cross validation to provide prior service quality characteristics. J Electr Syst. 2024;20:284–292.

37. Leinonen T, Wong D, Wahab A, Nadarajah R, Kaisti M, Airola A. Empirical investigation of multi-source cross-validation in clinical machine learning. arXiv preprint. 2024. 10.48550/arXiv.2403.15012.

38. Liaw R, Liang E, Nishihara R, Moritz P, Gonzalez JE, Stoica I. Tune: a research platform for distributed model selection and training. arXiv preprint. 2018. 10.48550/arXiv.1807.05118

39. Hastie T, Tibshirani R, Friedman JH. The elements of statistical learning: data mining, inference, and prediction 2. New York: Springer; 2009.

40. Pamuji FY, Putri SDA. Komparasi metode SMOTE dan ADASYN untuk penanganan data tidak seimbang MultiClass. J Inform Polinema 2023;9:331–338. 10.33795/jip.v9i3.1330.

41. Imani M, Ghaderpour Z, Joudaki M, Beikmohammadi A. The impact of SMOTE and ADASYN on random forest and advanced gradient boosting techniques in telecom customer churn prediction. In: 2024 10th International Conference on Web Research (ICWR). IEEE Publications 2024:202–209. 10.1109/ICWR61162.2024.10533320.

42. Halim AM, Dwifebri M, Nhita F. Handling imbalanced data sets using SMOTE and ADASYN to improve classification performance of Ecoli Data sets. Build. 2023;5:246–253. 10.47065/bits.v5i1.3647.

43. Johnson JM, Khoshgoftaar TM. Survey on deep learning with class imbalance. J Big Data. 2019;6:1–54. https://journalofbigdata.springeropen.com/articles/10.1186/s40537-019-0192-5.

